# Predicting Time to Diabetes Diagnosis Using Random Survival Forests

**DOI:** 10.1101/2024.02.03.24302304

**Authors:** Priyonto Saha, Yacine Marouf, Hunter Pozzebon, Aziz Guergachi, Karim Keshavjee, Mohammad Noaeen, Zahra Shakeri

## Abstract

Type 2 Diabetes Mellitus (T2DM) is a chronic metabolic disorder with increasing population incidence. However, T2DM takes years to develop, allowing onset prediction and prevention to be a clinically effective treatment strategy. In this study we propose and assess a novel approach to diabetes prediction which integrates a specialized extension of the random forest algorithm known as random survival forest (RSF). Rather than predicting a binary outcome, this machine learning model incorporates survival analysis methodology to predict the time until a patient will receive a diabetes diagnosis if their current lifestyle is maintained. We trained a baseline model on 7,704 electronic medical records from the Canadian Primary Care Sentinel Surveillance Network (CPCSSN) with 14 biomarker and comorbidity features across different measurement dates. Although tuning parameters were purposefully chosen for quick training rather than for predictive performance, our model exceeded expectations with a concordance index of 0.84. Thus, RSF models have been shown to produce accurate timelines of diabetes onset trajectory, providing patients with quantifiable and relatable risks that are easy to understand. The results of our study have substantial implications for advancing machine learning in clinical decision support and patient outcome predictions, emphasizing the role of innovative models in improving predictive accuracy.

## 1. INTRODUCTION

Type 2 Diabetes Mellitus (T2DM) is a chronic metabolic disorder where patients have difficulty controlling blood glucose levels due to insulin insensitivity [1]. As of 2019, 8.8% of Canadians are living with diabetes and there are approximately 549 new diagnoses daily [2]. Type 2 diabetes is incurable, so prevention and delaying onset is the best defense against this growing epidemic [3, 4]. The predominant methods of prevention include lifestyle and diet adjustments [5], and multiple studies report significant correlations between metabolic biomarkers, exercise rates, and diabetes incidence [6, 7].

Prior studies attempting to predict diabetes using Electronic medical records (EMR) often use traditional machine learning models that do not capture longitudinal measurements [**Samsel2024PredictingDepression**, 8–13]. Furthermore, studies using models with temporal features do not properly adjust for potential right censoring, that is, when a patient may develop diabetes after the study concludes.

Survival analysis methodology such as joint models and landmarking can address the right censoring prevalent in time-to-event data by predicting time to diabetes diagnosis. However, these techniques are often computationally intensive, make assumptions regarding underlying distributions, or struggle with analytical complexity [15].

To address these concerns, we propose the random survival forest (RSF) model, which is an ensemble method of survival trees to account for time to event data [16]. RSF is nonparametric, can utilize multiple longitudinal factors found in EMR data, and was found to outperform both joint models and landmarking [17]. By providing explicit time to diabetes diagnosis estimates, our model allows clinicians to better advise patients on their diabetes risk and collaborate towards personalized prevention plans for patients, thereby enhancing the efficacy of patient care strategies.

## II. Methods

### A. Data Collection and Preparation

The Canadian Primary Care Sentinel Surveillance Network (CPCSSN) provided a dataset of EMRs consisting of a random sample of 10,000 records with 43 features from 8,602 unique adult patients for model training. The dataset was generated by compiling systolic blood pressure (sBP) measurements with the closest in time clinical measurements from within a certain time frame. Clinical measurements of body mass index (BMI), low-density lipoprotein (LDL), high-density lipoprotein (HDL), total cholesterol (TC), and triglyceride (TG) were within one year, HbA1c (*A*1*c*) was within three months, and fasting blood sugar (*FBS*) was within one month. The dataset also included age and sex of each patient alongside binary comorbidity indicators of depression, hypertension (HTN), osteoarthritis (OA), and chronic obstructive pulmonary disease (COPD). Dates for all clinical measurements and health condition diagnoses were also included, with clinical measurement dates ranging between 2003 and 2015 and health conditions dates ranging from 1989 to 2015.

In the preliminary data exploration phase, we conducted an analysis of missing data, computed summary statistics, and generated a correlation matrix to assess the relationships between variables. We employed histograms to examine the distribution of predictor variables and utilized boxplots to identify potential outliers. This suite of visualizations was instrumental in uncovering patterns, anomalies, errors,and imbalances in the dataset, facilitating a more informed preprocessing strategy.

Survival time was calculated by taking the difference between the earliest date of a lab test or comorbidity diagnosis and their diabetes diagnosis date. The end of the study date, June 30th, 2015, was used for right-censored records where diabetes onset went unobserved. Left-censored individuals were removed, resulting in 7,756 eligible records with 14 predictors for the final dataset.

To identify the types of missing data, a series of logistic regression models was generated with the Statsmodels package[18]. For each feature, a binary missingness indicator was used as the response with all other features as covariates. Type of missingness was deemed missing completely at random (MCAR) if the logistic regression showed no significant covariate associations and missing at random (MAR) otherwise. Records where the only missing values were MCAR were dropped, and the remaining MAR missing data was imputed by chained equations through `IterativeImputer` from Scikit-Learn [19].

### B. Random Survival Forest Model

The random survival forest model is an extension of the random forest model that accounts for the presence of censoring in survival data. It uses the log-rank splitting rule that splits nodes by maximization of the log-rank test statistic [16, 20]. The survival forest model was implemented through scikit-survival library in Python 3.10.12 [19, 21].

To assess RSF models’ usefulness in predicting diabetes diagnosis timing, tuning parameters were selected for rapid training over optimal performance. This baseline random survival forest was trained with a 75:25 train-test split of our EMR data, consisting of 100 survival trees each with a max depth of 15, a minimum leaf sample of 100, and a minimum leaf split of 150.

To ensure replicability and facilitate further research, the source code of all the presented machine learning models is available on GitHub^1^

## III. Results and Discussion

### A. Preliminary Results

The initial exploratory analysis revealed no anomalies, with visualizations indicating that all clinical measures fell within expected ranges and exhibited no significant imbalances. The correlation matrix (Figure 1) showed no unexplained correlation. High correlation between total cholesterol, HDL, and LDL is explained by cholesterol in blood primarily consisting of HDL and LDL. Patients with higher *A*1*c* and *FBS* show higher rates of diabetes outcomes, which is expected since they are used as a diagnostic tool for diabetes [22]. Table I summarizes the 7,756 eligible records and their 14 non-chronological features before missing data processing, stratified on records with diabetes observed and diabetes unobserved. We determined that HDL and sBP were the only two features that were MCAR, which allowed us to drop records in which only these features were missing. LDL, TG, and total cholesterol were MAR and imputed, resulting in a sample size of 7,704.

**Fig. 1:**
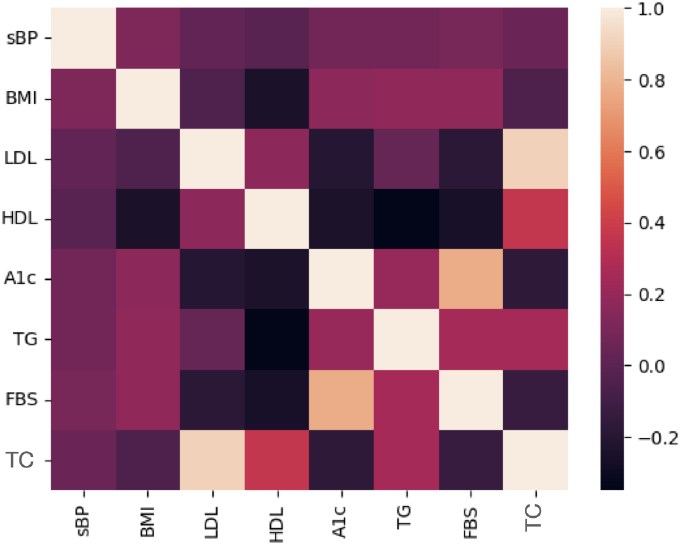
Correlation matrix of measured biomarkers. Zero represents no correlation and 1 represents perfect correlation.

**TABLE I:**
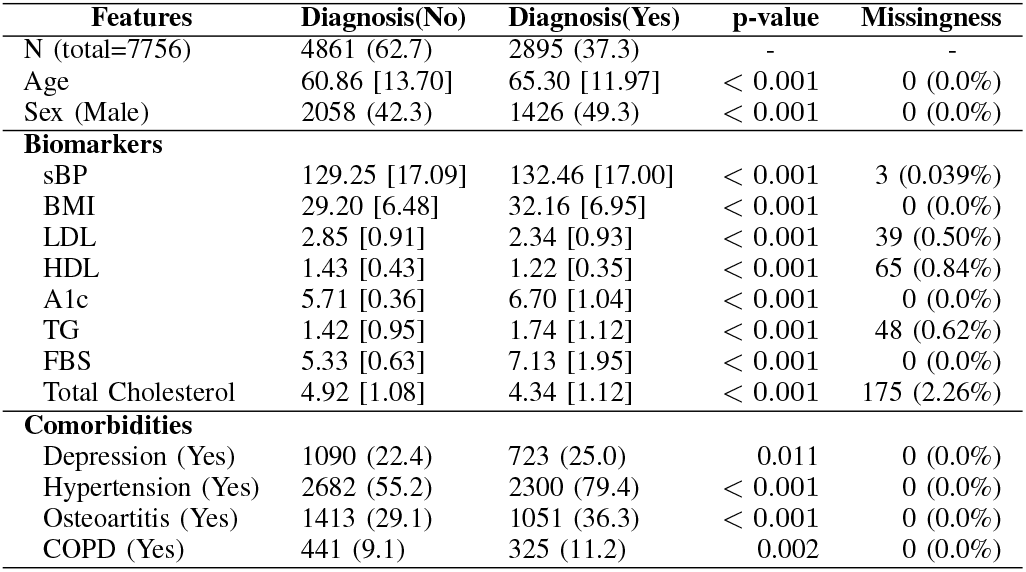
Summary statistics for 7,756 eligible records, stratified on records with diabetes observed and diabetes unobserved. Results are presented as mean [sd] for continuous variables or n (%) for categorical variables and missing values. P-values displayed are results of t-tests and chi-square tests for continuous and categorical features respectively

| Features | Diagnosis(No) | Diagnosis(Yes) | p-value | Missingness |
| --- | --- | --- | --- | --- |
| N (total=7756) | 4861 (62.7) | 2895 (37.3) | - | - |
| Age | 60.86 [13.70] | 65.30 [11.97] | < 0.001 | 0 (0.0%) |
| Sex (Male) | 2058 (42.3) | 1426 (49.3) | < 0.001 | 0 (0.0%) |
| **Biomarkers** |  |  |  |  |
| sBP | 129.25 [17.09] | 132.46 [17.00] | < 0.001 | 3 (0.039%) |
| BMI | 29.20 [6.48] | 32.16 [6.95] | < 0.001 | 0 (0.0%) |
| LDL | 2.85 [0.91] | 2.34 [0.93] | < 0.001 | 39 (0.50%) |
| HDL | 1.43 [0.43] | 1.22 [0.35] | < 0.001 | 65 (0.84%) |
| A1c | 5.71 [0.36] | 6.70 [1.04] | < 0.001 | 0 (0.0%) |
| TG | 1.42 [0.95] | 1.74 [1.12] | < 0.001 | 48 (0.62%) |
| FBS | 5.33 [0.63] | 7.13 [1.95] | < 0.001 | 0 (0.0%) |
| Total Cholesterol | 4.92 [1.08] | 4.34 [1.12] | < 0.001 | 175 (2.26%) |
| **Comorbidities** |  |  |  |  |
| Depression (Yes) | 1090 (22.4) | 723 (25.0) | 0.011 | 0 (0.0%) |
| Hypertension (Yes) | 2682 (55.2) | 2300 (79.4) | < 0.001 | 0 (0.0%) |
| Osteoarthritis (Yes) | 1413 (29.1) | 1051 (36.3) | < 0.001 | 0 (0.0%) |
| COPD (Yes) | 441 (9.1) | 325 (11.2) | 0.002 | 0 (0.0%) |

Our model had a concordance index (C-Index) of 0.84, which is the evaluation measure for random survival forest models [21]. This means that the model could accurately predict the relative order of diabetes diagnosis time of two randomly selected individuals 84% of the time [23].

Figure 2 shows the distribution of the median time of survival for diabetes diagnosis amongst records in the test set. In this context, the median time of survival is interpreted as the time before the model predicts that a record has over a 50% probability of diabetes diagnosis. Specifically, the bin into which a record is categorized signifies the estimated years until a patient is predicted to surpass a 50% probability of receiving a diabetes diagnosis. The median times for these records range from within one year to 15 years after the earliest biomarker measurement or health condition diagnosis date. Only 923 records of 1926 records in the test set are represented in the figure, with the remaining 1023 records predicted to never go above a 50% chance of diabetes diagnosis. Table II provides feature importance means, calculated using `permutation importance` from Scikit Learn [19, 21]. The most important model features were *A*1*c, FBS*, and total cholesterol, with feature importance means of 0.1009, 0.0734, and 0.00281, respectively. The standard deviations for these means were all less than 0.005. The importance of these features support current literature and diabetes monitoring standards [22].

**Fig. 2:**
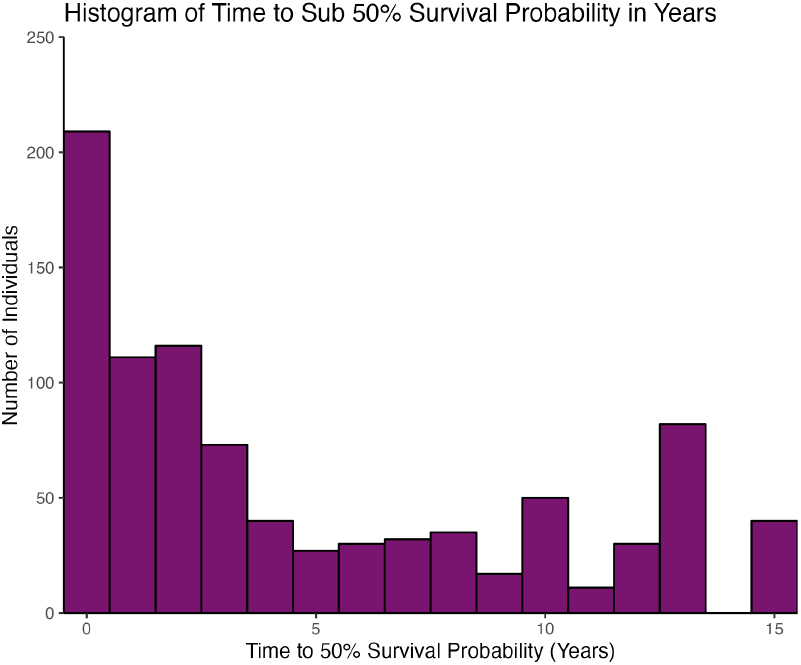
Median diabetes diagnosis time histogram for test set.

**TABLE II:** Feature Importance estimated with permutation.

| Features | Mean | Standard Deviation |
| --- | --- | --- |
| A1c | 0.12 | 0.0052 |
| FBS | 0.072 | 0.0050 |
| LDL | 0.0036 | 0.00087 |
| Total Cholesterol | 0.0030 | 0.00076 |
| BMI | 0.00079 | 0.00050 |
| Age at Exam | 0.00072 | 0.00026 |
| HTN | 0.00070 | 0.00044 |
| HDL | 0.00067 | 0.00030 |
| TG | 0.00055 | 0.00054 |
| Sex | 0.00011 | 0.000073 |
| COPD | 0.000045 | 0.000032 |
| Depression | 0.000030 | 0.000074 |
| OA | 0.000008 | 0.000075 |
| sBP | -0.000067 | 0.000099 |

### B. Key Findings

Overall, we found biomarkers and comorbidities commonly measured in health examinations to be important predictors for diabetes onset. To our knowledge, we are the first to utilize the RSF model to predict time to diabetes diagnosis through biomarkers and other comorbidites from a diverse Canadian population. Previous machine learning research has been conducted to predict diabetes, however they either utilize common random forests [24] or use RSFs for predicting diabetic complications [25]. One previous study employed the RSF model to predict the onset of diabetes, focusing on cardio-respiratory fitness and waistto-height ratio as key indicators [26]. This research was a prospective study confined to healthy male subjects, thus not encompassing a wider demographic spectrum.

#### Gender Diabetes Disparity

Based on Table I, men had a 32% higher likelihood of diabetes diagnosis compared to women. (OR: 1.32, 95%; CI: [1.20;1.45]). Gender differences in diabetes diagnosis are noteworthy, with Canadian census data showing higher prevalence among men. Notably, women in lower-income brackets experience even greater disparities in prevalence rates [27].

#### Diabetes Biomarker Differences

T-tests from Table I showed that *all* measured factors were significantly different between patients who became diabetic and patients who remained non-diabetic. On average, patients with diabetes had sBP that was in the healthy range (*<*130) whereas patients with diabetes had sBP in the high-normal range. Diabetic patients had higher BMI (*BMI >* 30) and differences in LDL, HDL, and total cholesterol compared to nondiabetics, but both groups remained within recommended ranges [28]. Diabetic patients exhibited borderline high TG levels, whereas non-diabetic patients maintained normal TG levels [29]. Average *A*1*c* and fasting blood glucose are also highly different between patients diagnosed with diabetes and healthy patients with varying health categories. Diabetes can be diagnosed through a fasting blood sugar measurement of over 7.0 mmol/L or through an A1c level above 6.5%. The average diabetic patient was above both those thresholds, while on average non-diabetic patients were well below the diabetic threshold and would not even fall within the prediabetic class, which requires a *FBS* between 6.1-6.9 mmol/L or an *A*1*c* of 6.0%6.4% for its diagnosis [30].

#### Enhanced RSF Prediction Granularity

Figure 2 elucidates the innovative aspect of our methodology; in contrast to conventional models that would categorize 923 records as *true* for a diabetes diagnosis, the RSF model provides a more nuanced differentiation. Predictions are rendered in days to facilitate precise and individualized care strategies, albeit aggregated into yearly intervals for histogram representation. As illustrated in Figure 2, the predominant segment of patients at risk are forecasted to manifest diabetes within five years from their initial measurement. Records not represented in the histogram, failing to surpass a 50% risk threshold, imply a low likelihood of these individuals developing diabetes in their lifespan. Owing to RSF’s capability to generate both a survival function and a cumulative hazard function per record, this inherent 50% benchmark can be adjusted to align with more conservative or liberal prognostic timelines. Therefore, the RSF model adeptly balances interpretability with comprehensive analytical depth, a challenge often encountered with alternative predictive models.

A C-Index of 0.84 indicates high predictive performance and shows great potential in the baseline capabilities of RSF. Furthermore, the simplicity of our pipeline leaves lots of room for optimization through parameter tuning. Although the flexible design of RSF was specifically proposed to accommodate the complexities of real world EMR, addressing data sparsity and imbalance provides another avenue for improvement. In particular for our dataset, the majority of patients had only a single record, multiple records were left-censored, and various records contained missing variables.

#### Longitudinal Data Challenges

Several advancements in random forest methodologies have been proposed to account for correlations among repeated measurements in longitudinal datasets [31]. Despite the potential of these enhancements to elevate predictive accuracy, our dataset’s absence of repeated measures precludes their application. In practical scenarios, Electronic Medical Records (EMR) from clinical settings are anticipated to offer an abundance of repeated measurements, thus paving the way for enhanced model refinement.

It is critical to distinguish between the dates of diagnosis and the actual onset of conditions, recognizing that patients may live with diabetes for a period before it is formally diagnosed. This scenario typifies interval censoring, a common challenge in survival analysis. The exact duration of this interval remains elusive in our study due to the prevalence of single-record patients, leading our model to estimate the time to diagnosis rather than the onset. Nonetheless, given the complications and potential emergencies arising from unmanaged diabetes, we posit that the interval between onset and diagnosis is likely minimal.

Looking ahead, our future work will concentrate on data generation processes to further refine our model’s framework and assess the effectiveness of random survival forest model extensions. This endeavor aims to optimize predictive performance and applicability in real-world settings.

## IV Conclusion

Diabetes is a lifelong condition that may greatly reduce one’s quality of life. The best defense against this growing epidemic is proactive prevention planning through lifestyle changes. However, implementing and maintaining these lifestyle changes can be challenging, especially without a solid understanding of the risks. This study advocates the use of random survival forest to predict the time to diabetes diagnosis, offering a quantifiable and relatable risk that is easy to comprehend. With a concordance index of 0.84 at baseline, random survival forest models have proven to be effective and accessible as a clinical tool with significant potential for further improvement. The complexities observed in our data reflect real-world constraints in electronic medical records. The random survival forest model excels under these conditions due to its robust non-parametric design. With such a promising initial performance, we are eager to explore additional applications of random survival forest in other prognostic modeling tasks.

## Data Availability

All data used are available online at https://cpcssn.ca/ website upon reasonable request.

## Footnotes

1 https://github.com/P-Saha/RSF-Diabetes-Prevention

